# Effects of the Mindfulness-Based Blood Pressure Reduction (MB-BP) Program on Depression and Neural Structural Connectivity

**DOI:** 10.1101/2021.08.18.21262240

**Authors:** Justin J. Polcari, Ryan J. Cali, Benjamin C. Nephew, Senbao Lu, Mikhail Rashkovskii, Julianne Wu, Frances Saadeh, Eric Loucks, Jean A. King

**Affiliations:** Dept. of Biology and Biotechnology, Worcester Polytechnic Institute, Worcester, Massachusetts; Dept. of Neurology, Massachusetts General Hospital and Harvard Medical School, Boston, Massachusetts; Dept. of Computer Science, Worcester Polytechnic Institute, Worcester, Massachusetts; Dept. of Behavioral and Social Sciences, Brown University, Providence, Rhode Island

## Abstract

Hypertension-related illnesses are a leading cause of disability and death in the United States, where 46% of adults have hypertension and only half have it controlled. It is critical to reduce hypertension, and either new classes of interventions are required, or we need to develop enhanced approaches to improve medical regimen adherence. The Mindfulness-Based Blood Pressure Reduction program (MB-BP) is showing novel mechanisms and early evidence of efficacy, but the neural correlates are unknown. The objectives of this study were to identify structural neural correlates of MB-BP using diffusion tensor magnetic resonance imaging (DTI) and assess potential correlations with key clinical outcomes. In a subset of participants from a larger randomized controlled trial, MB-BP participants exhibited increased interoception and decreased depressive symptoms compared to controls. Analyses of DTI data revealed significant group differences in several white matter neural tracts associated with the limbic system and/or hypertension. Specific changes in neural structural connectivity were significantly associated with measures of blood pressure, depression anxiety and symptoms, mindfulness, and emotional regulation. It is concluded that MB-BP has extensive and substantial effects on brain structural connectivity which could mediate beneficial changes in depression, interoceptive awareness, blood pressure, and related measures in individuals with hypertension.

## Introduction

Hypertension (defined as systolic blood pressure ≥130 mmHg and/or diastolic blood ≥80 mmHg) is associated with increased risk of cardiovascular diseases and associated morbidity and mortality (Oparil et al. 2018). As one of the most common chronic medical conditions, hypertension affects more than one billion adults worldwide with a prevalence of 31.1% (1.39 billion) in 2010 (Mills, Stefanescu, and He 2020). Risk factors for hypertension include alcohol consumption, physical inactivity, and unhealthy diet (Mills, Stefanescu, and He 2020), as well as major depression and anxiety (Grimsrud et al. 2009). Anxiety and major depression associations with hypertension have been attributed to the experience of chronic stress and/or trauma and deficits in emotional regulation (Grimsrud et al. 2009). Lifestyle changes, including diet and exercise (Oparil et al. 2018) and mindfulness meditation (Loucks et al. 2019; Nejati et al. 2015; Lee et al. 2020), have evidence to be effective in reducing high blood pressure.

Mindfulness meditation is particularly effective at attenuating stress, depression, and blood pressure (Ponte Márquez et al. 2019; de Vibe et al. 2017; Lee et al. 2020). An earlier study reported significant improvements in stress reduction, emotional regulation, in systolic and diastolic blood pressure in a mindfulness intervention group compared to matched controls (Nejati et al. 2015). The Mindfulness-Based Blood Pressure Reduction (MB-BP) program developed by our group has been used to evaluate the role of self-regulation mechanisms in blood pressure regulation (Loucks et al. 2019; Nardi et al. 2020). A single arm clinical trial of MB-BP suggests it may improve blood pressure and self-regulation skills and behaviors (physical activity, diet) of participants with elevated blood pressure (Nardi et al. 2020; Loucks et al. 2019). Specific mindfulness attributes, including emotional regulation, self-awareness, and attention control may mediate the reduction of hypertension and improvements in overall cardiovascular health (Nardi et al. 2020; Loucks et al. 2019; Loucks et al. 2015).

The relationship between depression and hypertension has been a recent area of interest in mindfulness-based studies. A thorough meta-analysis of the prevalence of depression in patients with hypertension has determined that there are distinct features of depression in patients with hypertension compared to those without (Li et al. 2015). Depression increases the risk for uncontrolled hypertension, with significant correlations between depression and both systolic and diastolic blood pressure values (Rubio-Guerra et al. 2013). While mindfulness training may be effective as an intervention for hypertension, as well as depression, the neural substrates of its impact on cardiovascular health are unknown.

Magnetic resonance imaging, specifically anatomical and diffusion tensor imaging (DTI), is a valuable tool for identifying the effects of both hypertension and mindfulness interventions on brain structure. DTI is an effective method for detecting changes in the structure of the neural tracts that connect brain regions. DTI studies of hypertension have identified decreased fractional anisotropy and increased mean diffusivity in the splenium of the corpus callosum, indicative of impaired neural integrity of these white matter tracts (Gons et al. 2012). Hypertension also leads to neural tract damage in the fornix of the limbic system (Sabisz, Naumczyk, Marcinkowska, Graff, Gąsecki, et al. 2019), a key substrate of mindfulness interventions. Another structural connectivity study of mindfulness identified increased connectivity in the corticospinal tract (Luders et al. 2011) and increased thickness of the corpus callosum region in the brains of meditators compared to that of control groups has been observed (Luders et al. 2012a). However, there have been no studies of the neural correlates of mindfulness training in participants with elevated blood pressure.

The objective of the present study was to identify the neural correlates of MB-BP using DTI. We hypothesized this integrative mindfulness intervention would alter structural connectivity in the limbic system (critically implicated in the effects of mindfulness intervention (Tang, Hölzel, and Posner 2015; Jiang et al. 2021; Luders et al. 2012b) and hypertension (Sabisz, Naumczyk, Marcinkowska, Graff, Gąsecki, et al. 2019; Hannawi et al. 2018) associated white matter tracts compared with the control condition. Our second objective was to investigate hypothesized associations between any changes in neural structural connectivity and primary (blood pressure) and relevant secondary (interoception, emotional regulation, perceived stress, anxiety, depression) outcomes.

## Methods

### Study sample and setting description

Participants were recruited and assessed during 2018-2020. The most common sources of participants were flyer/recruitment cards distributed throughout Rhode Island and Massachusetts, referral from friends, family members, coworkers or other person, referral from a former or current participant, and from a primary care practitioner or other health care professional.

Inclusion criteria for the overall RCT were: (1) Hypertension/elevated blood pressure (SBP≥120 mmHg systolic or DBP ≥80 mmHg); (2) Able to speak, read, and write in English; (3) All adults (≥18 years of age), genders and racial/ethnic groups were eligible to be included.

Exclusion criteria were: (1) Current regular meditation practice (>once/week); (2) Serious medical illness precluding regular class attendance; (3) Current substance abuse suicidal ideation or eating disorder; and (4) History of bipolar or psychotic disorders or self-injurious behaviors. These participants were excluded following standard guidelines (Santorelli and Kabat-Zinn 2003) because of risk for disrupting group participation, requiring additional or specialized treatment beyond capacity of this study, or already participating in practices similar to the intervention. The study protocol was approved by the institutional review board at Brown University (protocol #1412001171) on September 3, 2015. Participants provided written informed consent.

The present study includes clinical and MRI data from the baseline and 3-month (following completion of the MB-BP or active control protocol) time points.

### Intervention description

This study adapted Mindfulness-Based Stress Reduction (MBSR) for participants with prehypertension/hypertension, creating MB-BP. Specifically, MB-BP is based on, and time-matched to, the standardized MBSR intervention described elsewhere (Santorelli et al. 2017; Kabat-Zinn 2013). It consists of an orientation session, eight 2.5-hour weekly group sessions, and a 7.5-hour one-day session, and is led by a qualified MBSR instructor with expertise in cardiovascular disease etiology, treatment, and prevention. MB-BP and MBSR contain similar instruction and practices in mindfulness meditation, and conversations about stress and coping. Students learn a range of mindfulness skills including body scan exercises, meditation and yoga. Home practice consists of practicing mindfulness skills for ≥45 min/day, 6 days/week.

Unique aspects of MB-BP are education on hypertension risk factors, hypertension health effects, and specific mindfulness modules focused on awareness of diet, physical activity, medication adherence, alcohol consumption, stress, and social support for behavior change. MB-BP builds a foundation of mindfulness skills (e.g., meditation, yoga, self-awareness, attention control, emotion regulation) through the MBSR curriculum. MB-BP directs those skills towards participants’ relationship with their risk factors for hypertension.

MB-BP participants have their blood pressure and hypertension risk factors assessed at baseline and are provided with this information during the orientation session. During this session, the importance of hypertension for health and mortality is described, along with hypertension risk factors. This phase aims to engage participants’ interest in hypertension risk factors and increase motivation for behavior change. MB-BP encourages participants to explore personal readiness for change in the different hypertension risk factors and explore utilizing mindfulness practices to engage with those risk factors that they choose to. Instructors hold twenty-minute one-on-one interviews with each participant at the beginning of the course to foster a relationship between the instructor and participant, identify reasons for participation, and pinpoint opportunities for the instructor to customize the course to individual participants.

The course focuses on different determinants of BP each week. However, four common themes exist across all BP determinants, including: (1) Awareness of thoughts, emotions and physical sensations particularly surrounding determinants of BP such as food overconsumption, sedentary activities, alcohol consumption, and antihypertensive medication adherence. (2) Craving, particularly for determinants of BP such as overconsumption of palatable foods (e.g. those high in salt or sugar), sedentary activities, and alcohol consumption. (3) The impact of bringing mindfulness to every moment, particularly in relation to BP determinants, recognizing that this present moment is influenced by prior moments, including what we ate, the physical activity we had, and the amount of alcohol consumed. Participants are trained to bring non-judgmental attention to the often short-term pleasures of overconsumption of foods, sedentary activities, heavy alcohol consumption, or not taking antihypertensive medications, as well as to bring non-judgmental attention to the longer term suffering associated with these activities. Through this process, participants are encouraged to reflect on whether behavioral choices provide more benefit or harm to their well-being, and to choose beneficial behaviors. (4) Self-compassion: as self-regulatory and self-awareness skills increase as a result of the mindfulness practices, the curriculum emphasizes that it is common for participants to start caring for themselves in kinder ways. It is a way of better knowing ourselves, and through knowing ourselves in each moment, we often want to care for ourselves in each moment. This may mean taking medication that will support health, or being more physically active, eating more healthily, or consuming alcohol in more moderate amounts. As a whole, the MB-BP program trains participants in mindfulness skills, and then supports them to apply those skills to determinants of BP most relevant in their lives. After the 8-week course is completed, the only curriculum offerings are optional bimonthly MB-BP community group meetings, one-day retreats three times per year, and website access to new bimonthly meditations and talks. The MB-BP intervention took place in classrooms at the Brown University School of Public Health, in Providence, RI.

### Control Group Description

The active control group provided participants with, and trained them in the use of, a validated home blood pressure monitor (Omron, Model PB786N), which has evidence in and of itself to potentially lower blood pressure, (Tucker et al. 2017; Cappuccio et al. 2004) and is beyond usual care at this time. Participants who had Stage 2 hypertension (blood pressure >140/90 mmHg) were offered to have their physicians notified, if not already being overseen for uncontrolled hypertension. For participants with uncontrolled hypertension who do not have a physician, we worked with participants to provide access within constraints of their health insurance. Participants receive an educational brochure from the American Heart Association entitled “Understanding and Controlling Your High Blood Pressure Brochure” (product code 50-1731). Participants in the control group were offered a referral to the study’s psychiatrist if anxiety, depression, or suicidal ideation levels at baseline or follow-up reached clinical levels on the Beck Anxiety Inventory or the Revised Centers for Epidemiologic Studies Depression (CESD-R) scale. These services were also provided if participants requested them, regardless of mental health scale levels. Participants in the control group were eligible to take the MB-BP program after their 6-month follow-up assessment.

### Key outcomes

#### Blood Pressure

Clinic blood pressure was evaluated according to American Heart Association guidelines. (Pickering et al. 2005) Blood pressure was assessed using a calibrated Omron HEM-705CPN automated BP monitor (Lake Forest, IL) with established validity (Coleman et al. 2006). Participants rested, seated for 5 minutes with arm at heart level and legs uncrossed prior to blood pressure assessment. Participants were instructed to refrain from caffeine consumption and physical activity at least 30 minutes prior to blood pressure assessment. Blood pressure was assessed during the initial “screening assessment”, and then again at least one week later for the “baseline assessment.” Only the baseline assessment was used in order to remove potential upward biases in blood pressure because of stress-induced novelty resulting from attending the research clinic for the first time. At each assessment, three blood pressure readings were obtained with 60 seconds duration between assessments. The mean of the second and third blood pressure readings was used for analyses.

#### Multidimensional Assessment of Interoceptive Awareness (MAIA)

The MAIA is a 32 item self-report measure with response options ranging on a 6-point Likert scale (0/never-5/Always). Psychometric tests indicate the scale maintains moderate internal consistency (α = >0.70), good model (Comparative Fit Index = 0.886) (Mehling et al. 2012). The MAIA is composed of eight individual scales, specifically Noticing (awareness of uncomfortable, comfortable, and neutral body sensations); Not Distracting (tendency not to ignore or distract oneself from sensations of pain or discomfort); Not-Worrying (tendency not to worry or experience emotional distress with sensations of pain or discomfort); Attention Regulation (ability to sustain and control attention to body sensations); Emotional Awareness (awareness of the connection between body sensations and emotional states); Self-Regulation (ability to regulate distress by attention to body sensations); Body Listening (active listening to the body for insight); and Trusting (experience of one’s body as safe and trustworthy) (Mehling et al. 2012; Mehling et al. 2009). In order to reduce issues of multiple statistical testing, the primary outcome was an overall MAIA summary scale consisting of the mean of all items (including reverse coding when indicated). Secondary analyses evaluated impacts of MB-BP on each individual MAIA scale.

#### Difficulties in Emotion Regulation Scale (DERS)

The Difficulties in Emotional Regulation Scale (DERS) is a 36-item measure evaluating an individual’s capacity to regulate their emotional state (Gratz and Roemer 2004). Questionnaire responses are rated on a 5-point Likert scale (1 = almost never, 5 = almost always) and higher scores indicate a decreased ability to emotionally regulate (Gratz and Roemer 2004). The DERS has high internal consistency (α = 0.93) and good test/re-test reliability in validity studies (Gratz and Roemer 2004).

#### Perceived Stress, Depression, and Anxiety

Perceived stress was assessed utilizing the 10-item Perceived Stress Scale (PSS-10) with established validity and reliability (Roberti, Harrington, and Storch 2006; Cohen, Kamarck, and Mermelstein 1983). Participants were monitored for potential psychological distress (i.e. anxiety, depression and suicidal ideation) using the Beck Anxiety Inventory (BAI) and the Center for Epidemiology Study Depression Scale Revised (CESD-R) self-report questionnaires at each in-person research assessment.

### DTI Methods

Subjects were scanned on a Phillips (Phillips Medical Systems, Amsterdam, Netherlands) 3.0 Tesla Ingenia CX dStream MRI scanner at the University of Massachusetts Medical School Campus. Imaging data were acquired using a 3T MRI scanner (Phillips HealthCare Systems, Best, Netherlands). Axial diffusion weighted images were acquired using a spin echo DWiSE sequence. Scanning parameters were: acquisition matrix = 128 × 128, voxel size = 2.2 × 1.4 × 2.2mm^3, number of slices = 72. There were 63 directions acquired for the diffusion imaging (b = 1000 s/mm^2), including the b0 image (b = 0 s/mm^ 2). The repetition time was approximately 4000.00ms. The entire diffusion scan duration was 4 minutes and 30 seconds (270 seconds).

### Diffusion Tensor Image Analysis

Diffusion tensor imaging shows the intensity of images which is expressed by the rate of water in a specific gradient direction in brain regions. Using MRI, diffusion coefficients of brain regions can be determined (Sabisz, Naumczyk, Marcinkowska, Graff, Gąsecki, et al. 2019). These include: fractional anisotropy (FA) an overall measure of neural tract integrity, mean diffusivity (MD) an average value of total diffusion in a region of tissue, axial diffusivity (AD) which measures the magnitude of diffusion along a neural fiber tract and is often used as an index of axonal injury, and radial diffusivity (RD), the diffusion coefficient of flow perpendicular to neural fiber direction, which is often associated with myelin damage (Sabisz, Naumczyk, Marcinkowska, Graff, Gąsecki, et al. 2019; Winklewski et al. 2018).

Diffusion tensor analysis was performed using the FMRIB at Oxford’s neuroimaging software, FSL. Within FSL, the DTIFIT tool was used to assess any changes in fractional anisotropy (FA), mean diffusivity (MD), radial diffusivity (RD), and axial diffusivity (AD). Information on the specific instructions to run DTIFIT can be found here: fsl.fmrib.ox.ac.uk/fsl/fslwiki/FDT/UserGuide#DTIFIT. All data were registered to the standard MNI-152 (Montreal Neurological Institute) space. Upon completion of DTIFIT, tract-based spatial statistics (TBSS) were performed within FSL to extract any between-group differences in diffusion values changes over time (3 months) in the 19 limbic and hypertension related DTI tracks in the Johns Hopkins DTI atlas (https://neurovault.org/collections/264/, Table 1).

**Table 1:**
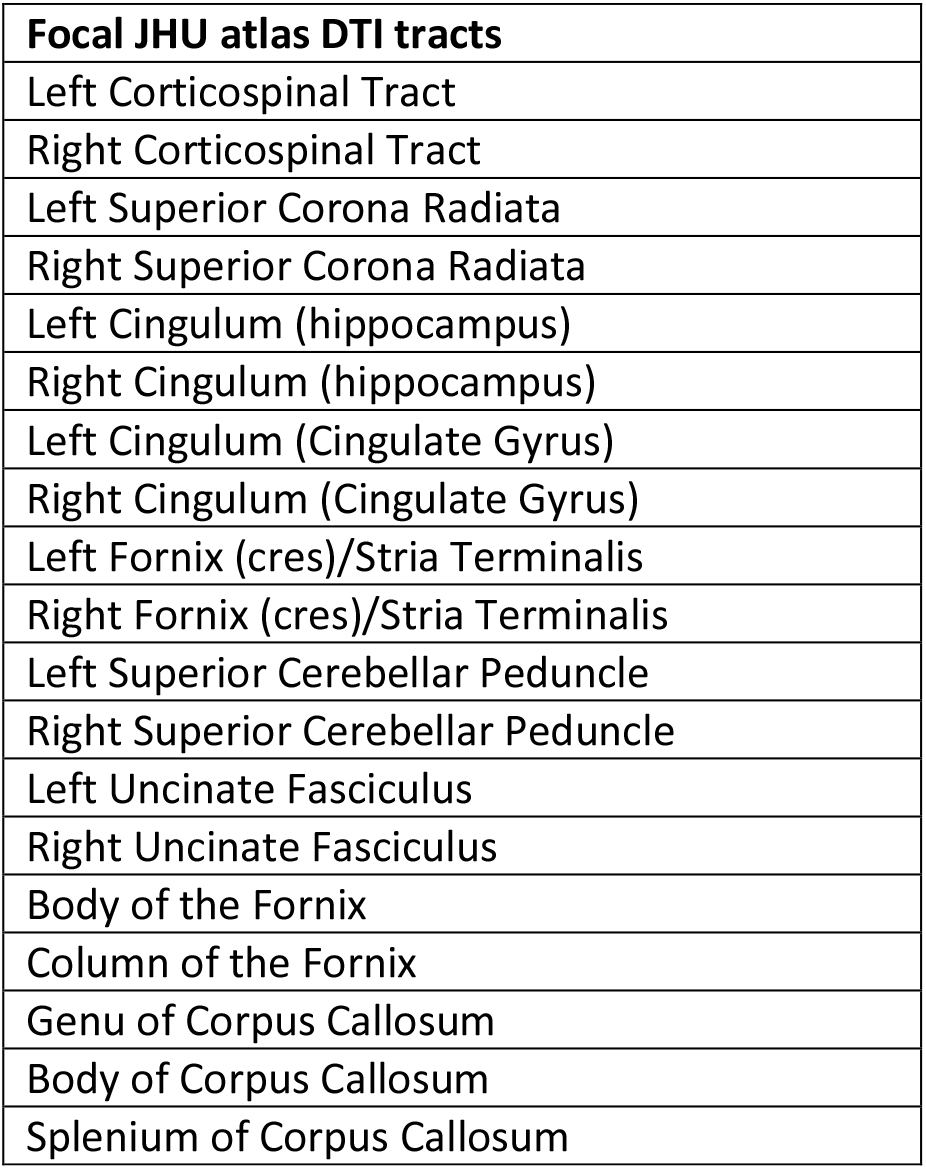
Limbic and hypertension associated DTI tracts from the Johns Hopkins University DTI Atlas.

### Correlations with Clinical Outcomes

Correlations between significant changes in DTI values and primary and secondary clinical outcomes were assessed using standard linear regression. There were no corrections for multiple comparisons due to the specific hypothesis driven focus on DTI tracts with significant group differences and selected clinical variables.

## Results

### Participants

In total, 36 eligible study participants completed pre- / post-fMRI imaging (14 MB-BP, 22 control, Table 1). Engagement with the intervention was measured using attendance data as well as daily home practice logs. The average adherence to the MB-BP intervention for those enrolled was 92%. Of the 14 participants assigned to the MB-BP intervention, only one withdrew from the intervention (at week 4). Attendance throughout the 8-week program remained high with 13 of the 14 enrolled having less than two absences. All thirteen attended the all-day retreat in full. Demographics show the study population (n = 36) included approximately 53% males and 47% females and was predominantly white race/ethnicity (72%; Table 2. Participants were highly educated on average with 78% earning a college degree or higher. The mean age for the control group was 57.0 years and for the MB-BP intervention group was 61.7 years (Table 2). Key factors included in the present analysis included: mean systolic blood pressure, mean diastolic blood pressure, interoceptive awareness (MAIA), emotional regulation (DERS), stress (PSS), anxiety (BAI), and depression (CESD) (Table 3). Using DTI MRI, statistically significant effects of MB-BP on changes over time of DTI variables in neural tracts were investigated from baseline to 3 month follow up (Table 4).

**Table 2:** Demographics of control and MB-BP intervention groups. The control group consisted of White (16), Black (2), Latino (2), Asian (1), and other (1). The MB-BP intervention group consisted of White (10), Black (3), and other (1). Education level, 4 = undergrad degree, 5 = graduate degree, 6 = postgraduate training.

| Clinical Demographic Data | Group | Mean ± Standard Error or % | T value | P value |
| --- | --- | --- | --- | --- |
| Gender | Control | 50.0% male, 50.0% female | 0.41 | 0.34 |
|  | MB-BP | 57.1% male, 42.9% female |  |  |
| Age, years | Control | 57.0 ± 3.4 | -0.99 | 0.16 |
| | MB-BP | 61.7 ± 2.7 | | |
| Education Level | Control | 4.8 ± 0.3 | -0.77 | 0.22 |
| | MB-BP | 5.2 ± 0.3 | | |

**Table 3:** Changes in key outcomes over 3 months in control and MB-BP groups.

| Clinical Data Change/Time | Group | Mean ± Standard Error | T value | P value |
| --- | --- | --- | --- | --- |
| Mean Systolic BP | Control | $-5.05 \pm 2.40$ | -0.22 | 0.42** |
| | MB-BP | $-4.00 \pm 4.72$ | | |
| Mean Diastolic BP | Control | $-4.52 \pm 1.59$ | -0.07 | 0.47** |
| | MB-BP | $-4.29 \pm 3.10$ | | |
| Interoceptive Awareness | Control | $-0.13 \pm 0.11$ | -5.23 | <0.001** |
| | MB-BP | $1.00 \pm 0.22$ | | |
| Emotional Regulation | Control | $1.55 \pm 2.18$ | 1.29 | 0.14** |
| | MB-BP | $-4.92 \pm 5.42$ | | |
| Perceived Stress | Control | $-0.59 \pm 0.80$ | 0.21 | 0.43** |
| | MB-BP | $-0.93 \pm 1.62$ | | |
| Anxiety | Control | $-0.05 \pm 0.69$ | 1.43 | 0.08 |
| | MB-BP | $-1.57 \pm 0.78$ | | |
| Depression | Control | $1.55 \pm 1.16$ | 1.75 | 0.045 |
| | MB-BP | $-1.36 \pm 1.00$ | | |

\*\*unequal variance t-test

**Table 4:**
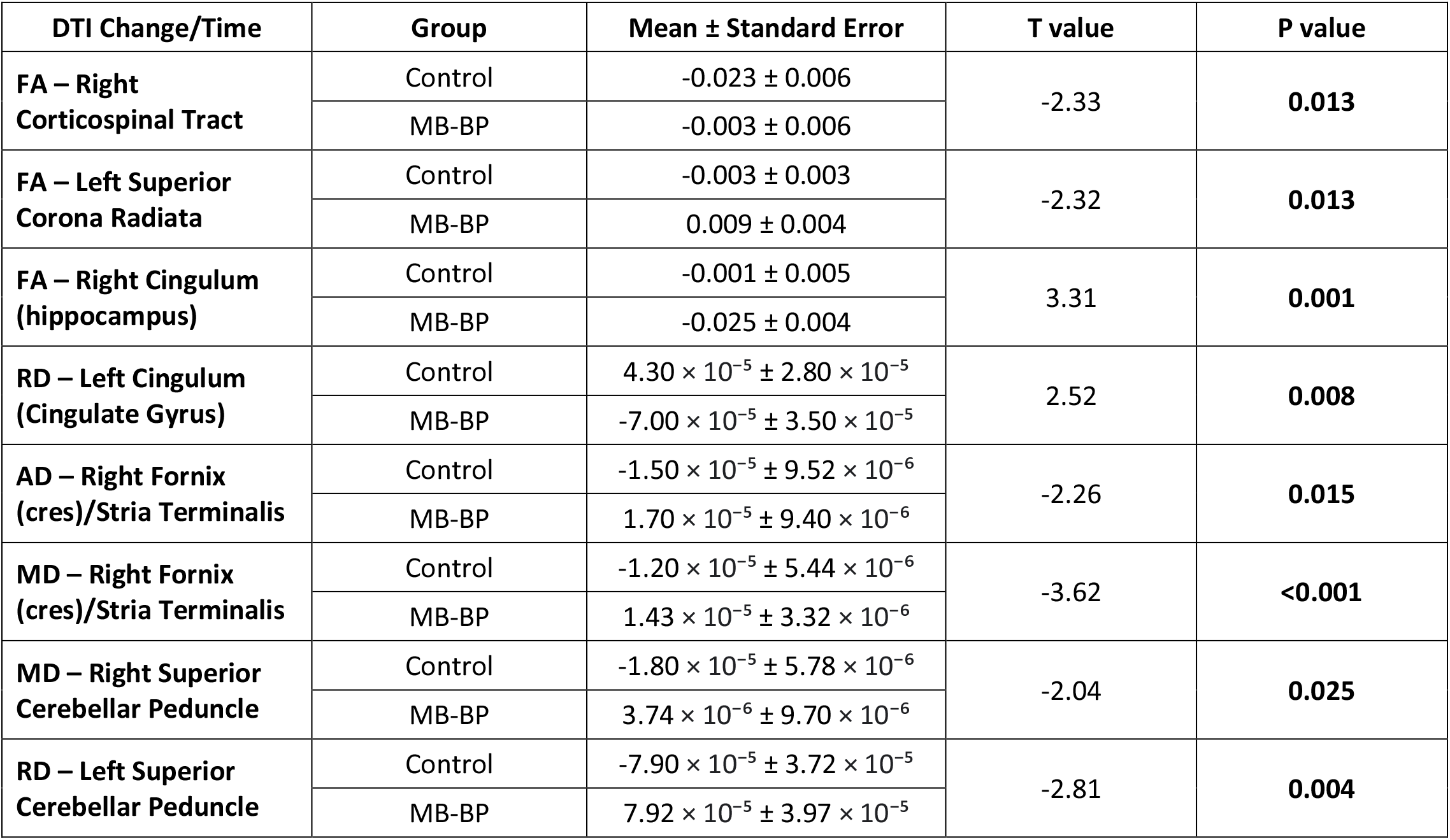
Mean changes in DTI metrics over 3 months in control and MB-BP groups. P-values represent between-group comparisons.

### Group differences in key outcomes

There were no significant differences in baseline values (p > 0.05). There were group differences in changes over time in interoceptive awareness (p < 0.001) and depression symptomology (p = 0.045), where interoception increased and depression symptoms decreased in the MB-BP group (table 3). In contrast, interoception was virtually unchanged and depression symptomology score increased in the control group. The interoceptive awareness score was 2.55 ± 0.17 at baseline and 2.42 ± 0.19 at 3 months in the control group (5.1% decrease) and 2.12 ± 0.18 at baseline and 3.12 ± 0.19 at 3 months (47% increase) in the MB-BP group (means ± SE, figure 1A). The depression symptomology score was 5.68 ± 0.94 at baseline and 7.23 ± 1.43 at 3 months in the control group (27% increase) and 4.29 ± 1.15 at baseline and 2.93 ± 1.26 at 3 months (32% decrease) in the MB-BP group (means ± SE, figure 1B). Given the comorbidity of depression and anxiety and role of anxiety in hypertension (Pan et al. 2015), there is a notable non-significant trend of a group difference in change over time of anxiety (p = 0.08), with anxiety decreasing in the MB-BP group. There were no statistically significantly group differences in changes in mean systolic blood pressure, mean diastolic blood pressure, emotional regulation, or perceived stress from baseline to 3 month follow up (table 3), although the adequately powered analyses in the full RCT sample found significant improvements in systolic blood pressure in MB-BP vs. control at the 6-month (but not 3-month) follow-up, suggesting effects on blood pressure reduction may take longer than the follow-up period for this imaging study.

**Figure 1A+B:**
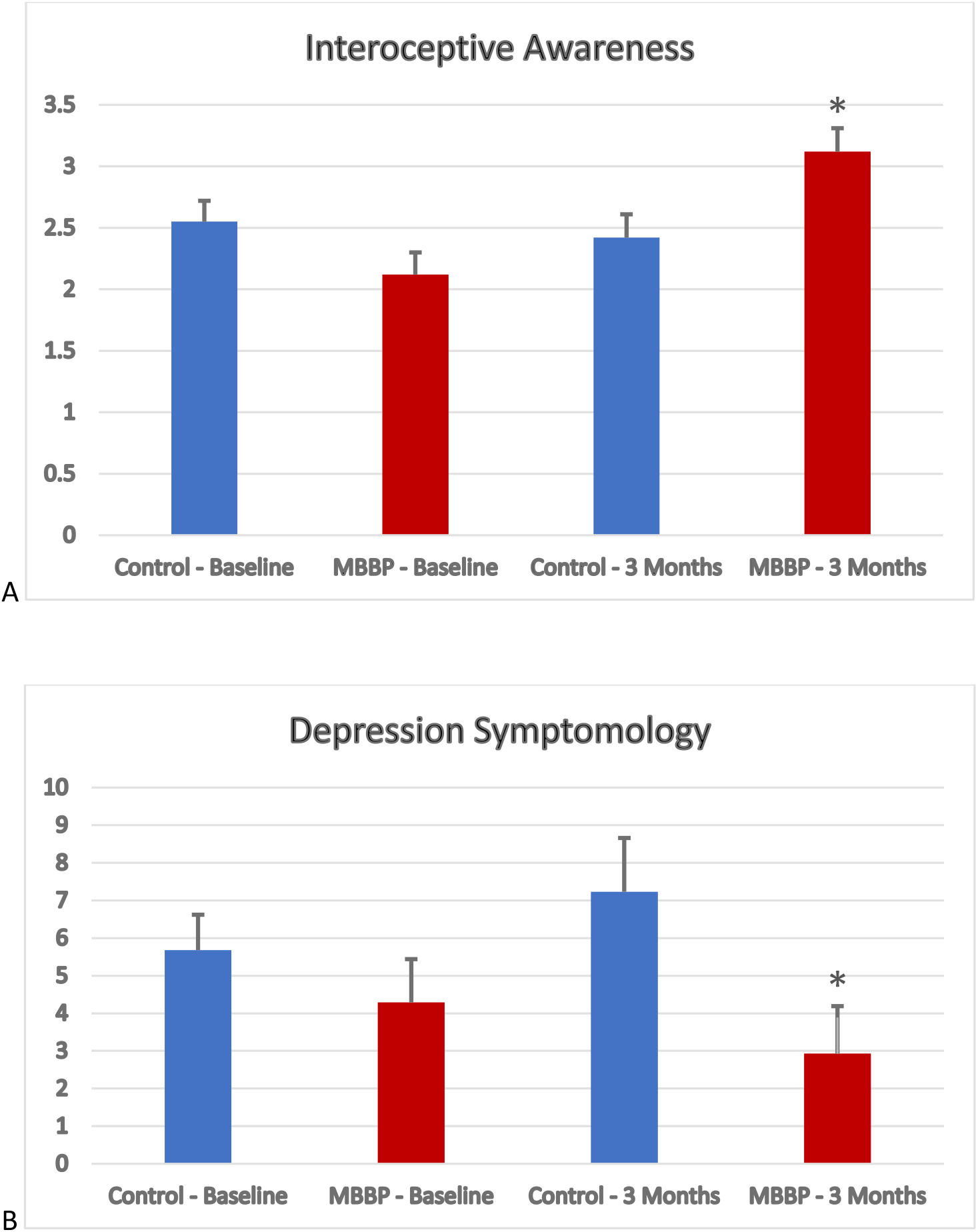
Mean interceptive awareness (MAIA, A) and depression symptomology (CESD, B) scores + standard error assessed at baseline and 3-months for control and MB-BP groups. * denotes significant group differences in change over time (p<0.05).

### Group Differences in DTI metrics

There were significant differences between MB-BP and control changes over time (baseline to 3 months) in the DTI metrics of several neural tracts (Table 4, Figure 2). Group differences in FA were identified in the right corticospinal tract (p = 0.013), left superior corona radiata (p = 0.013), and the right cingulum (hippocampus) (p = 0.001). RD changes differed in the left cingulum (cingulate gyrus) (p = 0.008) and the left superior cerebellar peduncle (p = 0.004). AD differed in the right fornix (cres)/stria terminalis (p = 0.015), and group differences in MD were identified in the right superior cerebellar peduncle (p = 0.025) and right fornix (cres)/stria terminalis (p < 0.001).

**Figure 2:**
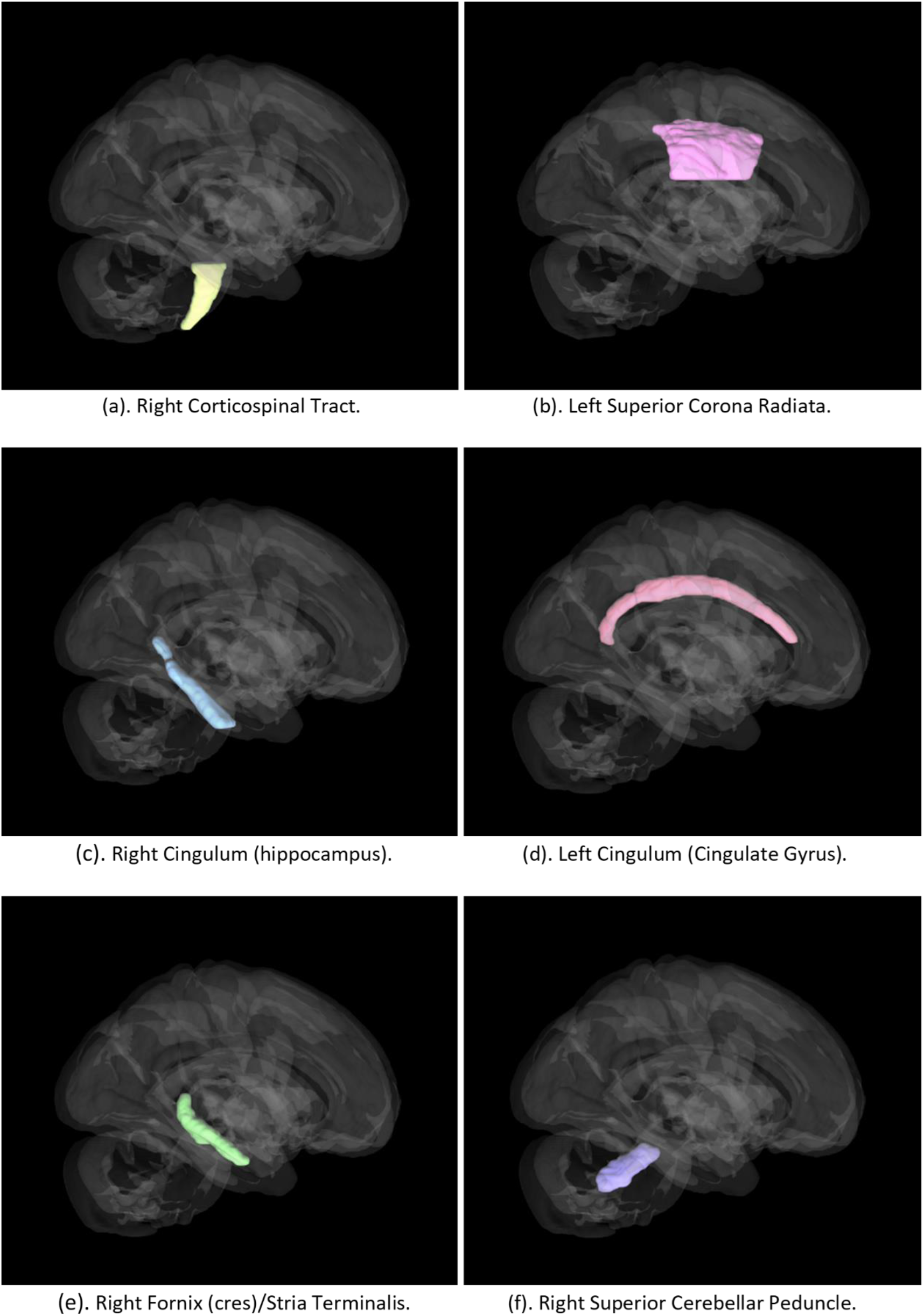
Locations of Johns Hopkins University atlas DTI tracts where there were significant group differences in DTI metric changes over time.

### Clinical outcome and DTI metric correlations

There were significant correlations between several clinical factors and DTI change/time data (Table 5). FA in the right corticospinal tract was significantly correlated with MAIA score (p = 0.032). AD in the right fornix (cres)/stria terminalis) tract was positively correlated with mean systolic blood pressure (p = 0.044), mean diastolic blood pressure (p = 0.010) and MAIA score (p = 0.005), and negatively correlated with depression symptomology (p = 0.011). MD in the right fornix (cres)/stria terminalis was positively correlated with MAIA (p = 0.002), and negatively correlated with depression symptomology (p = 0.011). RD in the left superior cerebellar peduncle was positively correlated with MAIA (p = 0.009) and negatively correlated with difficulty in emotional regulation (p = 0.003). RD in the left cingulum (cingulate gyrus), was negatively correlated with MAIA score (p = 0.027).

**Table 5:** Correlations between clinical factors and DTI metric changes over time, with significant correlations (bold) and trends with a white background.

|  | DTI Change/Time |  |  |  |  |  |  |  |
| --- | --- | --- | --- | --- | --- | --- | --- | --- |
|  | FA – Right Corticospinal Tract | FA – Left Superior Corona Radiata | FA – Right Cingulum (hippocampus) | AD – Right Fornix (cres)/Stria Terminalis | MD – Right Superior Cerebellar Peduncle | MD – Right Fornix (cres)/Stria Terminalis | RD – Left Superior Cerebellar Peduncle | RD – Left Cingulum (Cingulate Gyrus) |
| Mean Systolic BP | $R^2 = 0.016$<br>$r = 0.125$<br>$p = 0.464$ | $R^2 = 0.014$<br>$r = 0.117$<br>$p = 0.495$ | $R^2 = 0.0005$<br>$r = 0.022$<br>$p = 0.897$ | $R^2 = 0.114$<br>$r = 0.337$<br>$p = 0.044$ | $R^2 = 0.001$<br>$r = 0.031$<br>$p = 0.858$ | $R^2 = 0.066$<br>$r = 0.257$<br>$p = 0.130$ | $R^2 = 0.012$<br>$r = 0.107$<br>$p = 0.533$ | $R^2 = 0.054$<br>$r = -0.232$<br>$p = 0.174$ |
| Mean Diastolic BP | $R^2 = 0.046$<br>$r = 0.215$<br>$p = 0.206$ | $R^2 = 0.010$<br>$r = -0.101$<br>$p = 0.559$ | $R^2 = 0.001$<br>$r = 0.035$<br>$p = 0.843$ | $R^2 = 0.180$<br>$r = 0.424$<br>$p = 0.010$ | $R^2 = 0.002$<br>$r = -0.048$<br>$p = 0.781$ | $R^2 = 0.091$<br>$r = 0.301$<br>$p = 0.075$ | $R^2 = 0.030$<br>$r = 0.174$<br>$p = 0.310$ | $R^2 = 0.064$<br>$r = -0.254$<br>$p = 0.135$ |
| Interoceptive Awareness | $R^2 = 0.128$<br>$r = 0.358$<br>$p = 0.032$ | $R^2 = 0.007$<br>$r = 0.084$<br>$p = 0.627$ | $R^2 = 0.076$<br>$r = -0.275$<br>$p = 0.104$ | $R^2 = 0.210$<br>$r = 0.458$<br>$p = 0.005$ | $R^2 = 0.033$<br>$r = 0.180$<br>$p = 0.292$ | $R^2 = 0.258$<br>$r = 0.508$<br>$p = 0.002$ | $R^2 = 0.184$<br>$r = 0.429$<br>$p = 0.009$ | $R^2 = 0.136$<br>$r = -0.369$<br>$p = 0.027$ |
| Emotional Regulation | $R^2 = 0.070$<br>$r = -0.264$<br>$p = 0.124$ | $R^2 = 0.005$<br>$r = 0.072$<br>$p = 0.684$ | $R^2 = 0.0008$<br>$r = -0.028$<br>$p = 0.874$ | $R^2 = 0.083$<br>$r = -0.288$<br>$p = 0.093$ | $R^2 = 0.036$<br>$r = -0.189$<br>$p = 0.275$ | $R^2 = 0.042$<br>$r = -0.206$<br>$p = 0.235$ | $R^2 = 0.236$<br>$r = -0.486$<br>$p = 0.003$ | $R^2 = 0.042$<br>$r = 0.205$<br>$p = 0.239$ |
| Perceived Stress | $R^2 = 0.043$<br>$r = -0.207$<br>$p = 0.223$ | $R^2 = 0.002$<br>$r = -0.047$<br>$p = 0.789$ | $R^2 = 0.005$<br>$r = -0.072$<br>$p = 0.677$ | $R^2 = 0.052$<br>$r = -0.227$<br>$p = 0.183$ | $R^2 = 0.005$<br>$r = 0.074$<br>$p = 0.670$ | $R^2 = 0.001$<br>$r = -0.036$<br>$p = 0.835$ | $R^2 = 0.040$<br>$r = -0.199$<br>$p = 0.246$ | $R^2 = 0.098$<br>$r = 0.313$<br>$p = 0.063$ |
| Anxiety | $R^2 = 0.081$<br>$r = -0.285$<br>$p = 0.093$ | $R^2 = 0.061$<br>$r = -0.246$<br>$p = 0.148$ | $R^2 = 0.059$<br>$r = 0.242$<br>$p = 0.153$ | $R^2 = 0.024$<br>$r = 0.156$<br>$p = 0.364$ | $R^2 = 0.004$<br>$r = 0.060$<br>$p = 0.728$ | $R^2 = 0.029$<br>$r = 0.170$<br>$p = 0.320$ | $R^2 = 0.025$<br>$r = -0.157$<br>$p = 0.364$ | $R^2 = 0.016$<br>$r = 0.125$<br>$p = 0.464$ |
| Depression | $R^2 = 0.002$<br>$r = -0.040$<br>$p = 0.819$ | $R^2 = 0.036$<br>$r = -0.189$<br>$p = 0.271$ | $R^2 = 0.004$<br>$r = 0.062$<br>$p = 0.721$ | $R^2 = 0.178$<br>$r = -0.421$<br>$p = 0.010$ | $R^2 = 0.023$<br>$r = -0.150$<br>$p = 0.380$ | $R^2 = 0.175$<br>$r = -0.418$<br>$p = 0.011$ | $R^2 = 0$<br>$r = -0.005$<br>$p = 0.976$ | $R^2 = 0.041$<br>$r = 0.202$<br>$p = 0.238$ |

There were also three non-significant trends in the correlation data. There were trends for a negative correlation between AD in the right fornix (cres)/stria terminalis and difficulties with emotional regulation (p = 0.094), a positive correlation between MD in the right fornix (cres)/stria terminalis and mean diastolic blood pressure (p = 0.074), and a positive correlation between RD in the left cingulum (cingulate gyrus) and perceived stress (p = 0.063).

## Discussion

This study was conducted to complement and augment the stage 1 single arm clinical trial of MB-BP (Loucks et al. 2019) which showed significant improvements in blood pressure and stress, as well as the recently completed stage 2 randomized control trial. The present investigation identified increased interoceptive awareness, decreased depression symptoms and several group differences in changes over time in interoceptive awareness and/or hypertension associated white matter neural tracts in a subset of the MB-BP RCT subjects compared to controls. Specific changes in neural structural connectivity were significantly associated with measures of blood pressure, depression and anxiety symptoms, interoceptive awareness, and emotional regulation. It is concluded that MB-BP likely has extensive and substantial effects on brain structural connectivity which could mediate beneficial changes in depression and related clinical measures in individuals with hypertension.

The present clinical and interoception findings are consistent with the initial findings from the stage I single arm trial, where the novel, targeted MB-BP protocol was effective in enhancing interoceptive awareness (Loucks et al. 2019). MB-BP decreased depression symptoms in the current stage IIa population, and this finding underscores mindfulness’s efficacy with reducing depression (Takahashi et al. 2019). The substantial effect of MB-BP on depression symptomology is despite the presence of very low basal CESD scores in both groups. Individuals who participate in mindful awareness practices exhibit significant decreases in depression compared to individuals who participated in a health education program (Lopez-Maya, Olmstead, and Irwin 2019). Although not significant, we also observed a trend of decreased anxiety, which is often comorbid with depression (Pan et al. 2015). Prior investigation have identified similar effects, with mindfulness interventions inducing significant reductions in both depression and anxiety (Takahashi et al. 2019). While there were no group differences in measures of hypertension, this is likely due to lack of statistical power in the subset of participants enrolled in the imaging portion of the study compared to 48 participants in the stage 1 trial. The larger adequately power RCT, from which this sample is a subset, is showing significant improvements in systolic blood pressure for MB-BP vs. control at 6 months follow-up, that will be reported elsewhere (Loucks et al. In preparation.). In the RCT, effects on blood pressure were stronger at the later follow-up time point (i.e., 6 months) likely due to accumulating benefits from MB-BP’s impacts on health behavior change that can take time to exert effects on blood pressure. In addition to the increased interoceptive awareness and decreased depression symptoms, there were significant group differences in changes over time of several DTI measures in the targeted limbic and hypertension related white matter tracts.

The fornix is an arched shape bundle of nerve fibers that functions as the output tract of the hippocampus. It has major connections to the corpus callosum and lateral ventricles (Goldstein et al. 2021). Located in the mesial aspect of the cerebral hemispheres, the fornix connects multiple nodes of the limbic system and has essential roles in cognition and episodic memory recall (Senova et al. 2020). MD increased in the right fornix-stria terminalis tract in MB-BP subjects compared to controls, and this was driven by the increase in AD in this tract, indicating enhanced structural connectivity. Previous mindfulness imaging has reported thicker callosal regions in meditators, indicating greater structural connectivity (Luders et al. 2012a). This investigation was conducted using a multimodal imaging approach with diffusion tensor imaging (DTI) and structural magnetic resonance imaging (MRI) in addition to the collection of callosal measurements of tract-specific fractional anisotropy (Luders et al. 2012a). More recent DTI studies have illustrated that hypertension could contribute to white matter impairment and damage in the fornix (Sabisz, Naumczyk, Marcinkowska, Graff, Gąsecki, et al. 2019). Studies of hypertension have also explored correlations between hypertension and the diffusivity in the splenium of the corpus callosum, where hypertension was associated with lower fractional anisotropic values and significantly higher mean diffusivity in the splenium (Gons et al. 2012). Hypertension is associated with decreases in white matter fractional anisotropy in the fornix as well as the stria terminalis (Williams et al. 2019; Hannawi et al. 2018; Sabisz, Naumczyk, Marcinkowska, Graff, Gąsecki, et al. 2019), and mindfulness-based programs exert beneficial effects on structural properties of the stria terminalis (Feliu-Soler et al. 2020). Individuals with fibromyalgia (who often suffer from depression) randomized to mindfulness-based stress reduction exhibit greater increases in pain-flexibility, which is associated with changes in stria terminalis grey matter (Feliu-Soler et al. 2020). It is postulated that fornix and stria terminalis related neural tracts may be primary targets for the beneficial effects of mindfulness on hypertension and related comorbidities.

Hypertension has also been associated with functions of the anterior cingulate cortex (Gianaros et al. 2005). In the present study, RD increased in the cingulum-cingulate tract in controls and decreased in MB-BP, suggesting a beneficial effect of MB-BP on white matter structure in this region and supporting a prior report that short-term meditative practices induce white matter changes in the cingulate, as well as the corona radiate (Tang et al. 2010). Mindfulness has been specifically linked to structural changes in the cingulate (Mioduszewski et al. 2020; Laneri et al. 2015), as changes in white matter properties have been reported in the posterior cingulate cortex in experienced meditators (Laneri et al. 2015; Kang et al. 2013), the anterior cingulate cortex in experienced meditators and smokers, and healthy populations participating in mindfulness interventions (Kang et al. 2013; Laneri et al. 2015; Tang, Tang, and Posner 2016). MB-BP may attenuate hypertension related degeneration in cingulate cortex associated neural tracts through neurogenesis or myelination related mechanisms.

The corticospinal tract—a motor pathway made of white matter that travels from the cerebral cortex to lower motor neurons and interneurons in the spinal cord—controls voluntary movements of skeletal muscle of the body. In MB-BP participants, FA of the right corticospinal tract decreased less over time compared to controls, indicating attenuated deterioration in this tract. Mindfulness mediation and the corticospinal tract have been the focus of a previous study which reported pronounced differences in structural connectivity in the corticospinal tract, with the largest group differences observed between meditators and controls (non-meditators) (Luders et al. 2011). Given the unique physical activity-based aspects of MB-BP, it is hypothesized that this change in the corticospinal tract was mediated by changes in activity level, but evaluating this requires future detailed assessment of activity levels.

The corona radiata is a highly cellular white matter sheet that radiates from the internal capsule to the cerebral cortex. It transfers information between cerebral cortex and brain stem, connecting both motor and sensory nerve pathways. MB-BP increased FA in the left superior corona radiata, indicating improved neural integrity, where there this value decreased in controls. In a less comprehensive intervention, the FA of the left superior corona radiata was significantly increased after 10 hours of Integrative Body-Mind Training, suggesting that this tract contributes to emotional regulation (Tang, Tang, and Posner 2016). Earlier studies have also described hypointense lesions with unknown causes in several individuals with a history of chronic hypertension, suggesting a relationship between the corona radiata and chronic hypertension (Chan et al. 1996). This understudied neural feature may mediate the effects of MB-BP on neural pathways involved in both sensory/motor control and emotional regulation.

The cingulum is a ring-shaped white matter tract extending from the cingulate gyrus to the entorhinal cortex which facilitates communication within the limbic system. It connects the orbital frontal cortices, corpus callosum, and temporal lobe and also connects the subcortical nuclei to the cingulate gyrus. Cingulum fibers on the cingulate gyrus are involved in emotion and behavior regulation, underscoring its importance for studying mechanisms of schizophrenia and depression, while the cingulum fibers close to the hippocampus are involved in memory and cognitive functions (Bubb, Metzler-Baddeley, and Aggleton 2018). There was a larger decrease in FA in the right cingulum hippocampus tract in MB-BP subjects. The directionality of the present results also contrasts with an increase of FA in cingulum (cingulate gyrus and hippocampus) in long-term meditators (Luders et al. 2011), and a DTI study of hypertensive individuals who were otherwise healthy reported white matter disruption in the cingulum (Hannawi et al. 2018). Further research is needed to determine the significance of specific directional changes in connectivity measures in this tract.

There were several significant correlations between key outcomes and DTI change/time data. There was a significant correlation between interoceptive awareness and the right corticospinal tract, which supports similar reports of enhanced structural connectivity in the corticospinal tract of meditators compared to that of the control group of non-meditators (Luders et al. 2011). Another study with a focus on daily mindful Quadrate Motor Training for 6 weeks identified increases in fractional anisotropy in the tracts related to sensorimotor and cognitive functions, including the corticospinal tract (Piervincenzi et al. 2017). Given the activity components of both MB-BP and the Quadroto paradigm, exploration of the role of changes in activity level is warranted.

Correlations between the fornix and mean systolic and diastolic blood pressure and mean diastolic blood pressure were identified. Hypertension can contribute to white matter impairment and damage in the fornix (Sabisz, Naumczyk, Marcinkowska, Graff, Gąsecki, et al. 2019) and it has also been linked with decreased white matter FA in the stria terminalis and the fornix (Hannawi et al. 2018). Changes in DTI metrics in the fornix and stria terminalis are also associated with mindfulness features and depression symptoms in the present study, as increased fornix AD was inversely correlated with difficulty in emotional regulation and depression scores. Increased fornix MD was also inversely correlated with depression scores, which was driven by enhanced AD. Mindfulness has a regulatory role on depressive symptoms, inducing elevated levels of positive emotions and self-acceptance, (Jimenez, Niles, and Park 2010). Other investigations have characterized mindfulness related white matter changes in limbic structures including the corpus callosum, which is directly connected to the fornix, where mindfulness led to significant reductions in depression-dejection (Tang et al. 2012). A recent meta-analysis of mindfulness based cognitive therapy concludes it is an effective treatment for depression (Goldberg et al. 2019). Given the structural expanse and significance of the fornix, it is possible that this tract may be a key hub in coordinating mindfulness induced depression related changes across the brain.

Correlations were also present between the left superior cerebellar peduncle and both MAIA scores and emotional regulation, which supports prior studies. Increased cerebellar peduncle RD was inversely correlated with difficulty in emotional regulation, indicating that attenuated structural connectivity in this tract is associated with improved emotional regulation. While there is limited MRI literature on this region, a DTI study that investigated the relationships between microstructure of the cerebellar peduncle and cognitive function determined that the cerebellar peduncle had specific cognitive roles in sustained attention and working memory (Takahashi et al. 2010). Sustained attention and working memory are two important aspects of mindfulness, and the cerebellar peduncle could mediate and/or moderate the efficacy of mindfulness through these cognitive roles. The cerebellum functions in the regulation of emotional states related to motor, cognitive, and social behaviors (Adamaszek et al. 2017), underscoring the role of this region in emotional regulation, where it may integrate the cognitive and emotional effects of mindfulness interventions. MAIA scores were negatively correlated with RD in the left cingulum (cingulate gyrus) tract, indicating that increased mindfulness is associated with improved structural connectivity in the cingulum, generally consistent with prior research. A fMRI study that used a Continuous Performance Test to investigate neural tracts related to sustained attention identified significant activation in the cingulate gyrus while participants were conducting various tasks (Salgado-Pineda et al. 2004). In a DTI study focusing on Zen meditators compared to healthy non-meditator controls, long-term meditation was associated with significantly higher myo-inositol in the cingulate gyrus, which acts to regulate numerous cellular functions and suggests that the cingulate gyrus is involved in mediating the functioning of the default mode network (Fayed et al. 2013). Other investigations into mindfulness using functional connectivity analysis revealed stronger coupling between the posterior cingulate, dorsal anterior cingulate, and dorsolateral prefrontal cortices in experienced meditators (Brewer et al. 2011). Stronger coupling in these regions suggest that meditators have increased self-monitoring, cognitive control, and decreased mind-wandering—all key aspects of mindfulness, and this functional change may be mediated by enhanced structural connectivity in associated neural tracts.

Limitations include small sample size (leading to insufficient power in the analysis of hypertension) and the study duration (3 months). It is possible that the effect of mindfulness on blood pressure is cumulative, and that significant group differences would be present at later time points. The main MRI limitation is resolution in areas of extensive neural tract crossings, as DTI can characterize only one fiber in a volumetric pixel; thus, data obtained from a region of crossing fibers oriented in different directions may be unclear and/or misleading, as microscopic information is averaged.

While MB-BP is an effective complimentary intervention for hypertension, the neural correlates were previously unexplored. The current study identified several changes in neural structural connectivity in a unique group of tracts implicated in the effects of mindfulness and/or hypertension, clearly indicating that MB-BP has extensive and substantial effects on brain structural connectivity. Many of these changes were associated with measures of blood pressure, depression and anxiety symptoms, mindfulness, and emotional regulation, supporting the hypothesis that this targeted mindfulness intervention induces clinically relevant changes in the structure of the brain over a span of 3 months.

## Data Availability

For data inquiries, please reach out to the corresponding author Benjamin C. Nephew.

## Trial Registration

NCT from clinicaltrials.gov: 03256890

This study was supported by the National Institutes of Health (NIH) Science of Behavior Change Common Fund Program through an award administered by the National Center for Complementary and Integrative Health (UH3AT009145).

